# Maternal Knowledge and Education-Priority Gaps in Preterm Infant Care in the Gaza Strip, Palestine: A Cross-Sectional Study

**DOI:** 10.64898/2026.05.12.26353051

**Authors:** Mohammed Abdeljawad, Ahmed Najim, Hamza Abdeljawad, Josie Rodgers, Reham Almukbel, Kinan Mokbel

## Abstract

**Purpose:** To assess maternal knowledge of preterm infant care in Gaza and identify clinically actionable education priorities in a resource-constrained neonatal setting.

**Methods:** A cross-sectional survey was conducted among 170 mothers of premature infants admitted to neonatal departments in four government hospitals. A 30-item interviewer-administered questionnaire assessed knowledge across thermoregulation, feeding, phototherapy, and infection and skin care. Bivariate analyses, ordinal logistic regression, adjusted predicted probabilities, and exploratory clinical-priority gap analyses were conducted.

**Results:** Overall knowledge was moderate, with a mean score of 64.1% (SD 22.3). Knowledge was classified as poor in 53 mothers (31.2%), good in 41 (24.1%), and excellent in 76 (44.7%). Knowledge differed across domains (*p*<0.001), with feeding weakest (53.6%) and infection and skin care strongest (73.8%). Not receiving specialist premature-care antenatal follow-up was independently associated with lower odds of higher knowledge (adjusted OR 0.34, 95% CI 0.15–0.80, *p*=0.013). Mothers without specialist follow-up also had a higher adjusted probability of poor knowledge than those who received it (37.4% vs 18.1%) and more clinical-priority gaps (IRR 1.28, 95% CI 1.04–1.57, *p*=0.019). Among the 10 lowest-scoring items, 110 mothers (64.7%) had five or more gaps.

**Conclusion:** Maternal knowledge was uneven, with clinically important gaps in practical care domains. Domain-specific education checklists may strengthen antenatal counselling, bedside teaching, and discharge preparation in similar constrained neonatal settings.

**What is Known:**

◼ Mothers of premature infants need clear practical guidance to support safe feeding, thermal care, infection prevention and discharge preparation.
◼ Parent education in neonatal units may be inconsistent, especially in resource-constrained settings.

**What is New:**

◼ Maternal knowledge was uneven, with the weakest areas in feeding, phototherapy safety, handling practices and umbilical cord care.
◼ The study identifies a practical set of education-priority gaps that could be used to structure antenatal counselling, bedside teaching and discharge preparation.

## Introduction

Preterm birth, defined by the World Health Organization as birth before 37 completed weeks of gestation, remains a major contributor to neonatal morbidity and mortality worldwide.[1] Millions of infants are born preterm each year, with substantial variation between countries and settings.[2] Preterm birth complications are also an important contributor to child mortality.[3] Premature infants are vulnerable to longer-term health and developmental consequences, including neurodevelopmental impairment, chronic lung disease and later cardiometabolic or renal risk.[4] Very preterm infants are also at increased risk of sensory impairment, including hearing loss.[5] Large cohort data show substantial morbidity and mortality among extremely preterm infants, including respiratory distress, necrotising enterocolitis and sepsis.[6] Prematurity also carries longer-term health and educational costs,[7] while severe intraventricular haemorrhage remains an important acute morbidity in preterm infants.[8]

In the Gaza Strip, Palestine, prematurity is particularly important. WHO has described childbirth in Gaza as especially dangerous, with mothers and newborns facing overburdened hospitals and limited healthcare support before and after birth.[9] UNICEF has reported that 23% of births in Gaza are preterm, with approximately 10,000 neonates requiring transfer to neonatal intensive care units annually.[10] This burden occurs alongside widespread poverty and conflict-related maternal and neonatal health challenges.[11,12]

The care of premature infants requires informed participation from mothers and families. WHO guidance on care of preterm or low-birth-weight infants emphasises family involvement, parental support and education as part of routine care.[13] Maternal knowledge is clinically relevant because it can shape feeding practices, thermoregulation, infection prevention, recognition of danger signs and engagement with treatments such as phototherapy. Practical and emotional support from neonatal nurses in neonatal intensive care can also help mothers participate more confidently in care.[14]

Evidence on maternal knowledge of premature infant care in the Gaza Strip remains limited. This study therefore assessed maternal knowledge across four practical care domains in four government hospitals, examined associated sociodemographic, obstetric and service-related factors, and identified clinically actionable education priorities for resource-constrained neonatal settings.

## Materials and Methods

### Study design and setting

This cross-sectional study was conducted between March 2022 and March 2023 in the neonatal departments of four government hospitals in the Gaza Strip, Palestine: Al-Shifa Medical Complex, Al-Aqsa Hospital, Nasser Hospital and Emirati Hospital. These hospitals provide neonatal intensive care services and receive premature infants from across the Gaza Strip.

### Study population and sampling

The target population comprised mothers who had delivered a premature infant admitted to one of the selected neonatal units during the data collection period. Proportional sample allocation was based on estimated annual premature infant admissions across the four hospitals (Online Resource 1, Supplementary Table S1). The required sample size was calculated using Epi Info, based on a 95% confidence level and 5% margin of error. Consecutive sampling was used until the required sample for each hospital was reached. Mothers were eligible if they had delivered a premature infant admitted to one of the selected hospitals and provided written informed consent. Mothers were excluded if their infant was not present in the neonatal unit at the time of interview.

### Data collection instrument

Data were collected using a researcher-designed, interviewer-administered questionnaire developed in English and translated into Arabic. It included sections on sociodemographic characteristics, obstetric factors, maternal health services, sources of information and maternal knowledge of premature infant care. The English and Arabic versions are provided in Online Resource 1.

The knowledge section included 30 items across four domains: thermoregulation, feeding, phototherapy, and infection and skin care. Face and content validity were assessed by seven specialists, and a pilot study assessed feasibility and clarity. Internal consistency was assessed using Cronbach’s alpha.

### Knowledge scoring and statistical analysis

Each correct knowledge response was scored 1 and each incorrect response 0. Total and domain scores were expressed as percentages. Overall knowledge was categorised as poor (<50%), good (50-70%) or excellent (>70%), following Al-Mukhtar and Abdulghani.[15]

Data were analysed using Stata/SE version 19.0. Categorical variables were summarised as frequencies and percentages; continuous variables as means and standard deviations, with medians and interquartile ranges where appropriate. Pearson correlations examined relationships between knowledge domains. Domain scores were compared using the Friedman test, followed by Bonferroni-adjusted Wilcoxon signed-rank tests. Pearson chi-square tests assessed bivariate associations with knowledge category, with Monte Carlo sensitivity checks where expected counts were small.

Ordinal logistic regression identified independent predictors of higher knowledge category. The proportional odds assumption was assessed before interpretation. Sensitivity and exploratory analyses included binary logistic regression for poor versus good/excellent knowledge, adjusted predicted probabilities, domain-specific logistic regression for poor feeding and phototherapy knowledge, defined as domain scores below 50%, linear regression using the continuous overall score, and Poisson regression with robust standard errors for the count of incorrect responses across the 10 lowest-scoring knowledge items, with results reported as incidence rate ratios (IRR).

### Ethical considerations

Ethical approval was granted by the Helsinki Committee (HC) for Ethical Approval, Palestinian Health Research Council (PHRC), Gaza Strip, Palestine (approval number PHRC/HC/1025/22; 07 Feb 2022) in accordance with the Declaration of Helsinki.

### Large language model assistance

ChatGPT was used to support manuscript language editing. The authors verified the final content and take full responsibility for the work.

## Results

### Participant characteristics

Of 172 questionnaires distributed, 170 were completed, giving a response rate of 98.8%. The mean maternal age was 28.8 years (SD 6.4), and 51.2% of mothers had a bachelor’s degree or higher (Table 1; Online Resource 1, Supplementary Table S2). Most mothers were housewives (74.7%), and 77.6% of families had a monthly income below 1973 New Israeli Shekels (Online Resource 1, Supplementary Table S2). Pregnancy-induced hypertension was reported by 45.3% of mothers, previous Caesarean section by 62.4%, and twin pregnancy by 14.1%. Caesarean section was the mode of delivery in 77.1% of index births, and mean gestational age was 34.71 weeks (SD 1.50) (Table 1; Online Resource 1, Supplementary Table S3).

**Table 1.** Sociodemographic and obstetric characteristics of participants

| Characteristic | Category | N (%) | Mean ± SD |
| --- | --- | --- | --- |
| Maternal age (years) | <25 | 50 (29.4) | 28.8 ± 6.4 |
|  | 25–30 | 58 (34.1) |  |
|  | >30 | 62 (36.5) |  |
| Maternal education | Primary | 4 (2.4) |  |
|  | Preparatory | 27 (15.9) |  |
|  | Secondary | 52 (30.6) |  |
|  | Bachelor's degree or higher | 87 (51.2) |  |
| Family monthly income (NIS) | <1973 | 132 (77.6) | 1669 ± 1595 |
|  | 1973–2470 | 4 (2.4) |  |
|  | >2470 | 34 (20.0) |  |
| Pregnancy-induced hypertension | Yes | 77 (45.3) |  |
| Previous Caesarean section | Yes | 106 (62.4) |  |
| Twin pregnancy | Yes | 24 (14.1) |  |
| Mode of delivery (index birth) | Caesarean section | 131 (77.1) |  |
|  | Normal vaginal delivery | 39 (22.9) |  |
| Gestational age (weeks) | <34 | 22 (12.9) | 34.71 ± 1.50 |
|  | 34–35 | 97 (57.1) |  |
|  | >35 | 51 (30.0) |  |

**Note.** NIS, New Israeli Shekel.

### Maternal health services and sources of information

Mothers had a mean of 4.99 antenatal visits (SD 3.69), but only 28.2% reported specialist antenatal follow-up related to premature infant care. A prenatal examination by a doctor was reported by 61.2%, and 30.8% of mothers with available data had been told how to prepare for preterm delivery (Online Resource 1, Supplementary Table S4). Only 30.0% received information about premature infant care during pregnancy, whereas 82.4% received information after delivery, mainly from nurses or midwives (Online Resource 1, Supplementary Table S5).

### Maternal knowledge scores and priority gaps

The mean overall knowledge score was 64.1% (SD 22.3), with knowledge classified as poor in 31.2%, good in 24.1% and excellent in 44.7% of mothers (Table 2). Infection and skin care had the highest domain score (73.8%), followed by thermoregulation (67.4%), phototherapy (60.6%) and feeding (53.6%). The overall scale showed good internal consistency (Cronbach’s α=0.901), while domain alphas ranged from 0.593 to 0.773 (Table 2; Online Resource 1, Supplementary Table S6).

**Table 2.** Domain-specific and overall maternal knowledge scores.

| Knowledge domain | Items | Mean ± SD (%) | Range (%) | Median (IQR) | Cronbach's $\alpha$ |
| --- | --- | --- | --- | --- | --- |
| Thermoregulation | 10 | 67.4 ± 24.4 | 0.0–100.0 | 70.0 (50.0–90.0) | 0.773 |
| Feeding | 8 | 53.6 ± 28.6 | 0.0–100.0 | 50.0 (25.0–75.0) | 0.730 |
| Phototherapy | 5 | 60.6 ± 32.7 | 0.0–100.0 | 70.0 (20.0–80.0) | 0.748 |
| Infection and skin care | 7 | 73.8 ± 20.7 | 0.0–100.0 | 71.4 (57.1–100.0) | 0.593 |
| Overall knowledge* | 30 | 64.1 ± 22.3 | 13.3–100.0 | 65.0 (43.3–83.3) | 0.901 |

**Note.** Scores are percentages of correct responses. Overall knowledge categories were poor in 53 mothers (31.2%), good in 41 (24.1%), and excellent in 76 (44.7%). IQR, interquartile range.

Knowledge differed significantly across domains (Friedman χ²=104.11, df=3, *p*<0.001). Pairwise Wilcoxon signed-rank tests with Bonferroni correction showed that feeding scores were significantly lower than thermoregulation, phototherapy, and infection and skin care, while infection and skin care scores were higher than the other domains (Online Resource 1, Supplementary Table S7). Item-level results showed practical education gaps in handling practices, feeding tolerance, nasogastric feeding, phototherapy safety and umbilical cord care (Online Resource 1, Supplementary Tables S8–S12). Across the 10 lowest-scoring items, mothers had a mean of 5.6 priority gaps (SD 2.7), and 64.7% had five or more gaps (Table 3).

**Table 3.** Clinical-priority education gaps based on the 10 lowest-scoring knowledge items.

| Education-priority area | Knowledge item | Correct N (%) | Incorrect/gap N (%) | Practical implication |
| --- | --- | --- | --- | --- |
| Thermoregulation | Premature babies should not be manipulated frequently | 48 (28.2) | 122 (71.8) | Reinforce safe handling and heat-loss prevention |
| Feeding | Not all premature babies can tolerate breastfeeding | 58 (34.1) | 112 (65.9) | Explain feeding readiness and clinical variation |
| Phototherapy | The baby's genitalia should be covered during phototherapy | 70 (41.2) | 100 (58.8) | Include phototherapy safety in parent teaching |
| Infection and skin care | The diaper should not cover the umbilical cord | 70 (41.2) | 100 (58.8) | Reinforce cord care and infection prevention |
| Feeding | Burping a premature baby during and after feeding is essential | 73 (42.9) | 97 (57.1) | Include practical feeding support before discharge |
| Feeding | Feeding through NGT should not be done by pushing milk through a syringe | 83 (48.8) | 87 (51.2) | Clarify safe NGT feeding principles |
| Feeding | Pacifiers may be used for NPO premature babies | 84 (49.4) | 86 (50.6) | Explain non-nutritive sucking and NPO care |
| Phototherapy | The baby's position should be changed frequently during phototherapy | 85 (50.0) | 85 (50.0) | Reinforce positioning during phototherapy |
| Infection and skin care | The umbilical cord should not be covered | 86 (50.6) | 84 (49.4) | Clarify cord exposure and hygiene |
| Feeding | A premature baby may be kept NPO | 89 (52.4) | 81 (47.6) | Explain why oral feeding may be temporarily withheld |

**Note.** Items are presented from lowest to highest proportion of correct responses. Incorrect responses across these 10 items were counted as clinical-priority education gaps. NGT, nasogastric tube; NPO, nil per os.

### Associations with knowledge category

In bivariate analysis, knowledge category was associated with previous premature infants (χ²=10.75, *p*=0.029), birth order (χ²=14.68, *p*=0.023) and number of antenatal visits (χ²=10.66, *p*=0.031). Specialist premature-care antenatal follow-up showed a borderline association with knowledge category (χ²=5.85, *p*=0.054) (Table 4). Sociodemographic variables, including maternal age, place of residence, maternal education, husband’s education, family income, maternal work status and husband’s work status, were not significantly associated with knowledge category in bivariate analyses. Monte Carlo chi-square sensitivity checks for selected bivariate associations did not materially change the interpretation (Online Resource 1, Supplementary Table S13). Monte Carlo chi-square sensitivity checks for selected bivariate associations did not materially change the interpretation; variables significant in the Pearson chi-square analyses remained significant, and borderline or non-significant associations remained so (Online Resource 1, Supplementary Table S13).

**Table 4.**
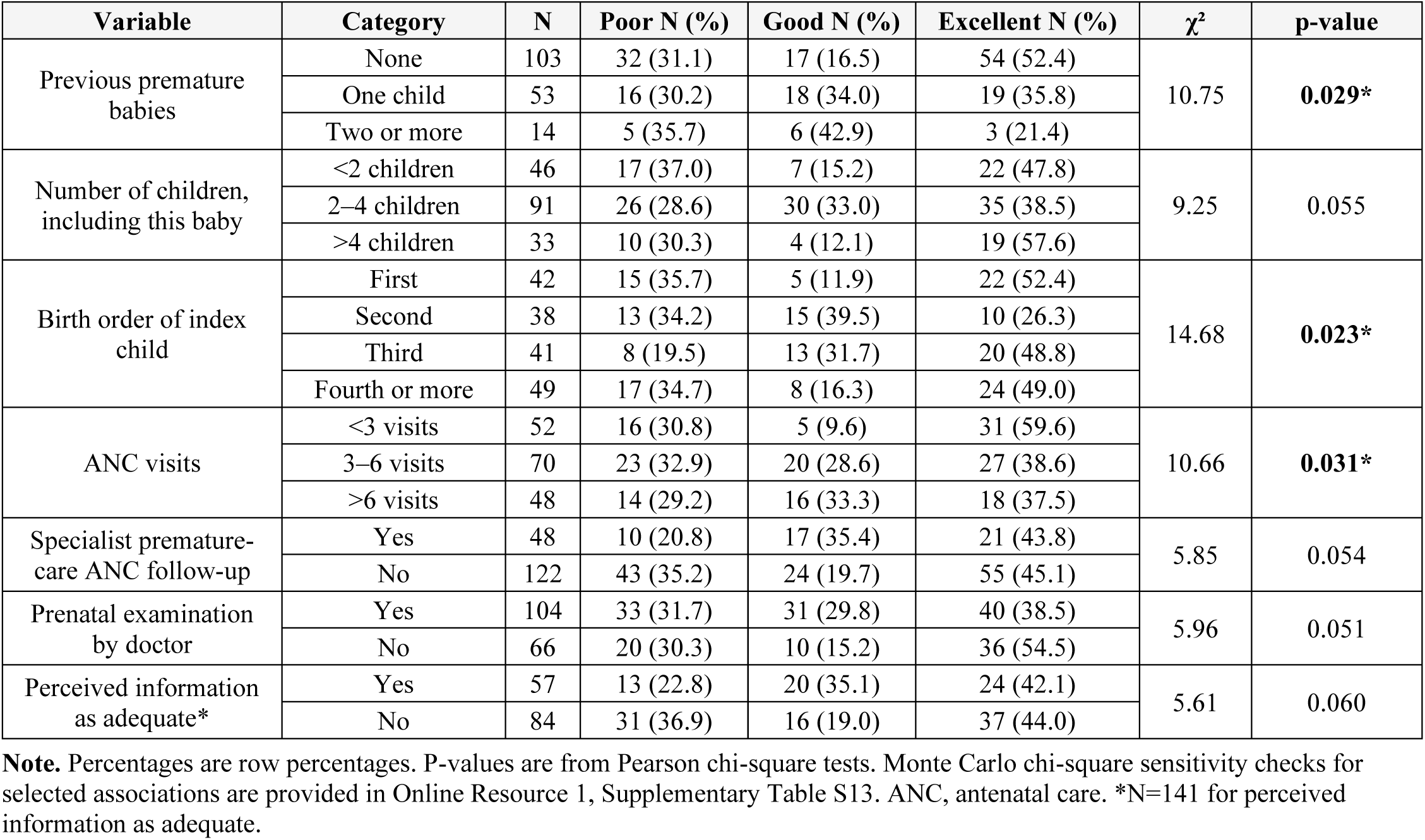
Bivariate associations between maternal knowledge category and selected variables.

| Variable | Category | N | Poor N (%) | Good N (%) | Excellent N (%) | $\chi^2$ | p-value |
| --- | --- | --- | --- | --- | --- | --- | --- |
| Previous premature babies | None | 103 | 32 (31.1) | 17 (16.5) | 54 (52.4) | 10.75 | **0.029\*** |
|  | One child | 53 | 16 (30.2) | 18 (34.0) | 19 (35.8) |  |  |
|  | Two or more | 14 | 5 (35.7) | 6 (42.9) | 3 (21.4) |  |  |
| Number of children, including this baby | <2 children | 46 | 17 (37.0) | 7 (15.2) | 22 (47.8) | 9.25 | 0.055 |
|  | 2–4 children | 91 | 26 (28.6) | 30 (33.0) | 35 (38.5) |  |  |
|  | >4 children | 33 | 10 (30.3) | 4 (12.1) | 19 (57.6) |  |  |
| Birth order of index child | First | 42 | 15 (35.7) | 5 (11.9) | 22 (52.4) | 14.68 | **0.023\*** |
|  | Second | 38 | 13 (34.2) | 15 (39.5) | 10 (26.3) |  |  |
|  | Third | 41 | 8 (19.5) | 13 (31.7) | 20 (48.8) |  |  |
|  | Fourth or more | 49 | 17 (34.7) | 8 (16.3) | 24 (49.0) |  |  |
| ANC visits | <3 visits | 52 | 16 (30.8) | 5 (9.6) | 31 (59.6) | 10.66 | **0.031\*** |
|  | 3–6 visits | 70 | 23 (32.9) | 20 (28.6) | 27 (38.6) |  |  |
|  | >6 visits | 48 | 14 (29.2) | 16 (33.3) | 18 (37.5) |  |  |
| Specialist premature-care ANC follow-up | Yes | 48 | 10 (20.8) | 17 (35.4) | 21 (43.8) | 5.85 | 0.054 |
|  | No | 122 | 43 (35.2) | 24 (19.7) | 55 (45.1) |  |  |
| Prenatal examination by doctor | Yes | 104 | 33 (31.7) | 31 (29.8) | 40 (38.5) | 5.96 | 0.051 |
|  | No | 66 | 20 (30.3) | 10 (15.2) | 36 (54.5) |  |  |
| Perceived information as adequate* | Yes | 57 | 13 (22.8) | 20 (35.1) | 24 (42.1) | 5.61 | 0.060 |
|  | No | 84 | 31 (36.9) | 16 (19.0) | 37 (44.0) |  |  |

**Note.** Percentages are row percentages. P-values are from Pearson chi-square tests. Monte Carlo chi-square sensitivity checks for selected associations are provided in Online Resource 1, Supplementary Table S13. ANC, antenatal care. \*N=141 for perceived information as adequate.

In adjusted ordinal logistic regression, not receiving specialist premature-care antenatal follow-up was independently associated with lower odds of higher knowledge category (adjusted OR 0.34, 95% CI 0.15-0.80, *p*=0.013). A higher category of previous premature infants was also associated with lower odds of higher knowledge (adjusted OR 0.51, 95% CI 0.29-0.88, *p*=0.016) (Table 5). The proportional odds assumption was not violated (global χ²=6.41, df=7, *p*=0.493). Binary logistic regression similarly showed that absence of specialist follow-up was associated with higher odds of poor knowledge (adjusted OR 3.07, 95% CI 1.11-8.46, *p*=0.030) (Online Resource 1, Supplementary Table S14).

**Table 5.** Adjusted ordinal logistic regression for predictors of higher maternal knowledge category (N=. 170)

| Predictor | Adjusted OR | 95% CI | p-value |
| --- | --- | --- | --- |
| Maternal education level, higher category | 1.07 | 0.71–1.63 | 0.732 |
| Previous premature infants, higher category | 0.51 | 0.29–0.88 | 0.016* |
| Birth order of index child, higher category | 1.22 | 0.91–1.64 | 0.175 |
| Number of ANC visits | 0.99 | 0.89–1.09 | 0.789 |
| No specialist premature-care ANC follow-up, vs yes | 0.34 | 0.15–0.80 | 0.013* |
| No prenatal doctor examination, vs yes | 2.28 | 0.99–5.26 | 0.054 |
| No information after delivery, vs yes | 1.34 | 0.58–3.08 | 0.494 |

Note. Outcome categories were ordered as poor, good, and excellent knowledge. OR >1 indicates higher odds of being in a higher knowledge category. For ordinal predictors labelled “higher category”, the OR represents the effect of a one-category increase. Nagelkerke pseudo- $R^2=0.085$ . The proportional odds assumption was not violated (global $\chi^2=6.41$ , $df=7$ , $p=0.493$ ). ANC, antenatal care; CI, confidence interval; OR, odds ratio.

Knowledge domains were positively correlated with each other (all *p*<0.001; Online Resource 1, Supplementary Table S15). Adjusted predicted probabilities also supported the main model: mothers without specialist follow-up had a higher predicted probability of poor knowledge than those with follow-up (37.4% vs 18.1%). A sensitivity analysis using the continuous overall knowledge score produced a similar pattern. A higher category of previous premature infants was associated with an 8.0 percentage-point lower overall knowledge score (95% CI −15.0 to −1.1, p=0.024), and absence of specialist premature-care antenatal follow-up was associated with an 11.5 percentage-point lower score (95% CI −21.6 to −1.4, *p*=0.025). Exploratory analyses showed that absence of specialist follow-up was associated with poor phototherapy knowledge (adjusted OR 2.95, 95% CI 1.14–7.59, *p*=0.025, lower continuous knowledge score and more clinical-priority gaps. Absence of specialist premature-care antenatal follow-up was associated with a higher number of priority gaps (IRR 1.28, 95% CI 1.04–1.57, *p*=0.019). A higher category of previous premature infants was also associated with more priority gaps (IRR 1.21, 95% CI 1.06–1.37, *p*=0.004). The association with no prenatal doctor examination was again counterintuitive and should be interpreted cautiously (RR 0.82,95% CI 0.67–1.00, *p*=0.049). Full exploratory and sensitivity analyses are shown in (Online Resource 1, Supplementary Table S16).

## Discussion

Previous studies have reported variable maternal knowledge or awareness of premature infant care. In Mosul, Iraq, Al-Mukhtar and Abdulghani reported that 53.0% of mothers had poor knowledge,[15] while a Rwandan study reported a mean awareness score of 59.3%.[16] In Gaza, Aldirawi et al. reported an overall post-discharge maternal knowledge value of 58.4%.[17] Abdullah and Hassan reported poor overall awareness of preterm baby home care among mothers in Erbil, Iraq.[18] These comparisons should be interpreted cautiously because the studies differed in sampling, questionnaire design, timing of assessment and scoring methods. The important finding in the present study is therefore not simply that knowledge was “moderate”, but that the gaps were concentrated in areas with immediate relevance to day-to-day care. A moderate total score can look reassuring, but it may conceal weaknesses in the specific tasks that mothers are expected to understand during admission and after discharge.

Infection and skin care was the strongest domain, consistent with previous Gaza-based findings.[17] This is encouraging, particularly because preterm infants are biologically vulnerable to infection and because hand hygiene remains central to neonatal infection prevention.[19,20] However, the domain-level score should not be interpreted as evidence that infection-prevention knowledge was fully adequate. Important gaps remained in hand hygiene and umbilical cord care. These are not minor details. They are routine caregiving behaviours that can be taught clearly, checked quickly and reinforced before discharge. The findings therefore suggest that even relatively stronger domains still need item-level review, because the mean score alone may miss clinically relevant weaknesses.

Feeding was the weakest domain. This aligns with evidence from Rwanda and Brazil showing that feeding and home-care knowledge can be challenging for mothers of premature infants.[16,21] This is a particularly important finding because feeding is one of the most demanding areas of premature infant care. Mothers may understand broad messages about the value of breast milk or frequent feeding, but still be uncertain about why some premature infants cannot tolerate direct breastfeeding, why oral feeding may be withheld, or why nasogastric feeding may be used. Optimising nutrition is central to the care of preterm and low-birth-weight infants.[22] Enteral nutrition in preterm infants is also complex and requires careful clinical guidance rather than generic infant-feeding advice.[23] From the parental perspective, feeding is shaped by mothers’ developing understanding of their infant’s needs and feeding cues.[24] Preterm infants have distinct nutritional requirements,[25] and post-discharge micronutrient supplementation remains an area where clearer consensus is needed.[26] This makes feeding a clear priority for structured parent education.

The thermoregulation and phototherapy findings also point to the need for more specific teaching. Mothers generally recognised some basic principles, but weaker items related to frequent handling, phototherapy protection and positioning suggest gaps in practical knowledge. Studies have reported gaps in maternal knowledge of thermoregulation and essential newborn care.[27,28] Maintaining appropriate body temperature remains clinically important in very-low-birth-weight infants.[29] Variable maternal or caregiver knowledge of neonatal jaundice has also been reported,[30,31] and clinical summaries emphasise the practical relevance of understanding jaundice management.[32] These areas are clinically important because parents may participate in routine care while the infant is still vulnerable. Education should therefore move beyond broad reassurance and use short, concrete messages, such as how to reduce heat loss during handling, why positioning matters during phototherapy and what protection is required during treatment.

The lowest-scoring item analysis strengthens the practical value of the study. The weakest items were not randomly distributed across the questionnaire. They clustered around feeding tolerance, nasogastric feeding, phototherapy safety, handling practices and umbilical cord care. These are precisely the areas where a short education checklist could be useful. Such a checklist would not need to replace clinical judgement or individualised advice. Rather, it could provide a minimum set of topics that should be addressed with every mother of a premature infant before discharge. In resource-constrained neonatal settings, this is important because education may be delivered under pressure, by different staff members and at different points in the care pathway.

The most actionable service-related finding was the association between specialist premature-care antenatal follow-up and higher maternal knowledge. Antenatal care has been linked with newborn-care knowledge in other settings.[33] WHO guidance also emphasises repeated antenatal contacts beginning early in pregnancy.[34] In the present study, only 28.2% of mothers reported specialist antenatal follow-up related to premature infant care, despite all participants ultimately delivering prematurely. This suggests a missed opportunity. Broader socioeconomic, psychosocial and food-security factors have also been associated with preterm labour, reinforcing the need to interpret antenatal education within the wider conditions in which mothers receive care.[35] Antenatal education research suggests that structured education can improve mothers’ knowledge of newborn care.[36] In Gaza, where health services operate under sustained pressure, strengthening the educational content of existing antenatal contacts may be more feasible than relying only on additional visits.[37]

Postnatal education remains important, but the adjusted analysis suggests that it should be interpreted cautiously. Most mothers received information after delivery, mainly from nurses and midwives, yet receipt of information after delivery was not independently associated with higher knowledge category. This does not mean that postnatal education is ineffective. A more plausible interpretation is that information may vary in content, timing, quality or retention. Previous work from Gaza and Brazil has emphasised the role of neonatal nurses in educating mothers of premature infants.[17,21] Studies of parental knowledge and parent-provider communication also suggest that communication in neonatal care needs to be structured, consistent and responsive to parents’ needs if it is to support understanding and participation.[38,39] Bedside teaching may therefore be most useful when it is standardised around key topics but still adapted to the infant’s clinical condition and the mother’s existing knowledge.

The role of non-clinical information sources should also be interpreted carefully. Very few mothers reported mass media as a source of information during pregnancy, which limits inference from this variable. Public health messaging may help raise awareness, but it cannot substitute for clinically grounded education on premature infant care. Parent-targeted postnatal education interventions in low- and middle-income countries vary in content, delivery method and outcomes, so their value depends on careful integration with clinical care.[40] For this study, the implication is clear: education should be delivered as part of the care pathway, not left to chance or informal sources.

Sociodemographic variables, including maternal education and family income, were not significantly associated with knowledge category in this study. This contrasts with studies from Iraq, India, Nepal and Ethiopia in which maternal education was associated with mothers’ knowledge of premature infant care, home-based neonatal care, newborn care or neonatal danger signs.[15,41,42,43] Several explanations are possible. The relatively high proportion of mothers with bachelor’s degrees or higher may have reduced variation in education within the sample. It is also possible that, in a highly constrained setting, access to targeted clinical information is more influential than general educational background. This interpretation is important because it challenges the assumption that more educated mothers necessarily have adequate knowledge of premature infant care. Education delivered by clinical teams remains necessary even when mothers have relatively high formal education.

A further important finding was that previous premature birth was independently associated with lower odds of higher knowledge and with more priority gaps. This is counterintuitive, because previous experience might be expected to improve knowledge. The finding should therefore be interpreted cautiously. Previous premature birth may identify mothers with more complex obstetric histories, repeated exposure to stressful neonatal care, or earlier missed opportunities for counselling. It may also reflect residual confounding. Even so, the practical message is valuable: previous experience of prematurity should not be treated as evidence of adequate preparation. Mothers with previous premature births may still need structured and individualised education.

This study has several strengths. It was conducted across multiple government hospitals, achieved a high response rate and examined knowledge both as an overall score and across clinically meaningful domains. The item-level analysis also improved clinical interpretability by identifying the specific areas where education should be prioritised. The statistical approach was strengthened by Monte Carlo sensitivity checks, assessment of the proportional odds assumption and exploratory analyses using alternative outcomes. These analyses supported the main finding that specialist premature-care antenatal follow-up was associated with better maternal knowledge.

The study also has limitations. Its cross-sectional design prevents causal inference, and the questionnaire was researcher-designed, which may limit comparability with other studies. Consecutive sampling may introduce selection bias, and the findings may not generalise to private hospitals, non-hospital settings or culturally different populations. Self-report and interviewer-administered responses may also be affected by recall or social desirability bias. More broadly, maternal experiences of caring for preterm infants are shaped by social vulnerability, family circumstances and health-system pressures.[44] The data were collected before the major escalation in Gaza from October 2023, and the current educational and healthcare context for mothers of premature infants may now be substantially more disrupted.

These findings support a practical response. Neonatal teams in resource-constrained settings could use a short domain-specific education checklist covering feeding tolerance, NPO and nasogastric feeding, phototherapy safety, safe handling and umbilical cord care. This would help ensure that the most clinically relevant gaps are addressed consistently during antenatal counselling, bedside education and discharge preparation. Future studies should test whether structured antenatal and postnatal education improves maternal knowledge, confidence, caregiving practices and infant outcomes, and whether remote support, such as video consultation, can support early in-home care after discharge.[45]

## Conclusion

This study found moderate maternal knowledge of premature infant care in the Gaza Strip, but with clear gaps in clinically important areas, especially feeding, phototherapy safety, handling practices and umbilical cord care. Specialist premature-care antenatal follow-up was associated with better knowledge and fewer priority gaps, while previous premature birth did not appear to provide adequate preparation. In resource-constrained neonatal settings, a short domain-specific education checklist may offer a practical way to strengthen antenatal counselling, bedside teaching and discharge preparation. Future studies should test whether structured education improves maternal knowledge, caregiving confidence and infant outcomes.

## List of Abbreviations

ANC: Antenatal care
NGT: Nasogastric tube
NIS: New Israeli Shekel
NPO: Nil per os
CI: Confidence interval
IRR: Incidence rate ratio
OR: Odds ratio
SD: Standard deviation

## Funding

This research received no specific grant from any funding agency in the public, commercial or not-for-profit sectors.

## Competing interests

The authors have no relevant financial or non-financial interests to disclose.

## Consent to participate

Written informed consent was obtained from all participants.

## Data Availability

De-identified participant data are available from the corresponding author upon reasonable request, subject to ethical approval and applicable data protection regulations.

## Acknowledgements

For the purpose of open access, the authors have applied a Creative Commons Attribution (CC BY) licence to any Author Accepted Manuscript version arising from this submission.

## Author Contributions

MA: data collection, analysis and interpretation. AN and HA: study conception and design and revised the manuscript for intellectual content. JR contributed to drafting and critical revision of the manuscript and contributed to data interpretation. RA: contributed to statistical analysis, data interpretation and critically revised the clinical aspects of manuscript. KM provided senior oversight, led manuscript development, contributed to the analysis and interpretation of data and critically revised it. All authors approved the final version of the manuscript and agree to be accountable for all aspects of the work.

## Notes

### Competing Interest Statement

The authors have declared no competing interest.

### Funding Statement

This study did not receive any funding

