## Supplementary materials for "Maternal Knowledge and Education-Priority Gaps in Preterm Infant Care in the Gaza Strip, Palestine: A Cross-Sectional Study"

This file contains Supplementary Tables S1-S16 and the English and Arabic versions of the study questionnaire.

#### Appendix 1. Supplementary tables

**Supplementary Table S1.** Proportionate sample size by hospital

| Hospital | Estimated admissions/year | Estimated admissions/2 months | Required sample |
|---|---|---|---|
| Al-Shifa Medical Complex | 996 | 166 | 100 |
| Nasser Hospital | 437 | 72 | 43 |
| Emirati Hospital | 147 | 24 | 15 |
| Al-Aqsa Hospital | 128 | 21 | 14 |
| Total | 1708 | 283 | 172 |

**Note.** These figures were based on hospital admission estimates and were used only for proportional sample allocation; they were not participant-level data.

**Supplementary Table S2.** Full sociodemographic characteristics of participants (N=170)

| Characteristic | Category | N (%) | Mean ± SD |
|---|---|---|---|
| Maternal age (years) | <25 | 50 (29.4) | 28.8 ± 6.4 |
| | 25–30 | 58 (34.1) | |
| | >30 | 62 (36.5) | |
| Place of residence | Rafah | 23 (13.5) | |
| | Khan Younes | 38 (22.4) | |
| | Middle Gaza | 20 (11.8) | |
| | Gaza | 66 (38.8) | |
| | North Gaza | 23 (13.5) | |
| Maternal education | Primary | 4 (2.4) | |
| | Preparatory | 27 (15.9) | |
| | Secondary | 52 (30.6) | |
| | Bachelor's degree or more | 87 (51.2) | |
| Husband's education | Primary | 4 (2.4) | |
| | Preparatory | 12 (7.1) | |
| | Secondary | 52 (30.6) | |
| | Bachelor's degree or more | 102 (60.0) | |
| Family monthly income (NIS) | <1973 | 132 (77.6) | 1669 ± 1595 |
| | 1973–2470 | 4 (2.4) | |
| | >2470 | 34 (20.0) | |

| | | |
|---|---|---|
| Mother's work status | Employed/worker | 43 (25.3) |
| | Housewife | 127 (74.7) |
| Husband's work status | Employed/worker | 133 (78.2) |
| | Not working | 37 (21.8) |

**Note:** The term 'husband' reflects the wording used in the locally reviewed questionnaire and was considered contextually appropriate by the expert panel during questionnaire development.

**Supplementary Table S3.** Obstetric characteristics of participants (N=170)

| Characteristic | Category | N (%) | Mean ± SD |
|---|---|---|---|
| Diabetes mellitus | Yes | 10 (5.9) | |
| | No | 160 (94.1) | |
| Pregnancy-induced hypertension | Yes | 77 (45.3) | |
| | No | 93 (54.7) | |
| Premature rupture of membranes | Yes | 8 (4.7) | |
| | No | 162 (95.3) | |
| Previous Caesarean section | Yes | 106 (62.4) | |
| | No | 64 (37.6) | |
| Twin pregnancy | Yes | 24 (14.1) | |
| | No | 146 (85.9) | |
| Mode of delivery | Normal vaginal delivery | 39 (22.9) | |
| | Caesarean section | 131 (77.1) | |
| Previous premature babies | None | 103 (60.6) | |
| | One child | 53 (31.2) | |
| | Two or more | 14 (8.2) | |
| Gestational age (weeks) | <34 | 22 (12.9) | 34.71 ± 1.50 |
| | 34–35 | 97 (57.1) | |
| | >35 | 51 (30.0) | |
| Number of children including this baby | <2 children | 46 (27.1) | 2.81 ± 1.74 |
| | 2–4 children | 91 (53.5) | |
| | >4 children | 33 (19.4) | |
| Birth order | First | 42 (24.7) | |
| | Second | 38 (22.4) | |
| | Third | 41 (24.1) | |
| | Fourth or more | 49 (28.8) | |
| Sex of baby | Male | 82 (48.2) | |
| | Female | 88 (51.8) | |

**Supplementary Table S4.** Maternal health services received (N=170 unless otherwise stated)

| Variable | Category | N (%) | Mean ± SD |
|---|---|---|---|
| Number of ANC visits | <3 visits | 52 (30.6) | 4.99 ± 3.69 |
| | 3–6 visits | 70 (41.2) | |
| | >6 visits | 48 (28.2) | |
| Specialist ANC follow-up related to premature care | Yes | 48 (28.2) | |
| | No | 122 (71.8) | |
| Prenatal examination by doctor | Yes | 104 (61.2) | |
| | No | 66 (38.8) | |
| Prenatal examination by nurse/midwife | Yes | 99 (58.2) | |
| | No | 71 (41.8) | |
| Told how to prepare for preterm delivery* | Yes | 52 (30.8) | |
| | No | 117 (69.2) | |

***N=169** for "told how to prepare for preterm delivery" because one response was missing.

**Supplementary Table S5.** Sources of information about premature infant care

| Variable | Category | N (%) | Denominator |
|---|---|---|---|
| Received information during pregnancy | Yes | 51 (30.0) | 170 |
| | No | 119 (70.0) | 170 |
| Doctor, during pregnancy | Yes | 43 (84.3) | 51 |
| Nurse/midwife, during pregnancy | Yes | 14 (27.5) | 51 |
| Mass media, during pregnancy | Yes | 3 (5.9) | 51 |
| Relative/friend, during pregnancy | Yes | 5 (9.8) | 51 |
| Received information after delivery | Yes | 140 (82.4) | 170 |
| | No | 30 (17.6) | 170 |
| Doctor, after delivery | Yes | 55 (39.3) | 140 |
| Nurse/midwife, after delivery | Yes | 124 (88.6) | 140 |
| Mass media, after delivery | Yes | 8 (5.7) | 140 |
| Relative/friend, after delivery | Yes | 22 (15.7) | 140 |
| Perceived information as adequate | Yes | 57 (40.4) | 141 |
| | No | 84 (59.6) | 141 |

**Supplementary Table S6.** Internal consistency of the knowledge scale

| Scale/domain | Number of items | Cronbach's α |
|---|---|---|
| Thermoregulation | 10 | 0.773 |
| Feeding | 8 | 0.730 |
| Phototherapy | 5 | 0.748 |
| Infection and skin care | 7 | 0.593 |
| Overall knowledge scale | 30 | 0.901 |

**Supplementary Table S7.** Comparison of knowledge scores across domains

| Comparison | Mean difference, percentage points | Median difference | Adjusted *p*-value |
|---|---|---|---|
| Thermoregulation vs feeding | 13.8 | 15.0 | <0.001 |
| Thermoregulation vs phototherapy | 6.8 | 10.0 | 0.002 |
| Thermoregulation vs infection and skin care | −6.4 | −5.7 | <0.001 |
| Feeding vs phototherapy | −7.0 | −5.0 | 0.005 |
| Feeding vs infection and skin care | −20.2 | −21.4 | <0.001 |
| Phototherapy vs infection and skin care | −13.2 | −17.1 | <0.001 |

**Note.** Domain scores were compared using the Friedman test, which showed a statistically significant difference across the four domains ($\chi^2$=104.11, df=3, p<0.001). Pairwise comparisons were conducted using Wilcoxon signed-rank tests with Bonferroni correction for multiple comparisons. Positive mean and median differences indicate higher scores in the first listed domain.

**Supplementary Table S8.** Thermoregulation knowledge items (N=170)

| Item | Correct N (%) | Incorrect N (%) | Rank |
|---|---|---|---|
| The premature baby should be placed in a warm environment | 106 (62.4) | 64 (37.6) | 5 |
| The premature baby loses body heat rapidly if uncovered | 91 (53.5) | 79 (46.5) | 9 |
| Covering a premature baby's head helps keep the baby's temperature | 142 (83.5) | 28 (16.5) | 4 |
| Premature babies should be placed near open windows with air drafts | 153 (90.0) | 17 (10.0) | 2 |
| A premature baby could be placed on cold beds or blankets | 95 (55.9) | 75 (44.1) | 8 |
| Premature babies should be manipulated frequently | 48 (28.2) | 122 (71.8) | 10 |
| A source of heat is essential to keep a baby's temperature normal | 96 (56.5) | 74 (43.5) | 7 |
| Handling the baby with warmed hands is necessary | 145 (85.3) | 25 (14.7) | 3 |
| Dressing the baby in previously warmed clothes is necessary | 106 (62.4) | 64 (37.6) | 6 |
| The baby should be bathed with warm water and immediately dried and covered | 164 (96.5) | 6 (3.5) | 1 |

**Supplementary Table S9.** Feeding knowledge items (N=170)

| Item | Correct N (%) | Incorrect N (%) | Rank |
|---|---|---|---|
| Any premature baby can tolerate breastfeeding | 58 (34.1) | 112 (65.9) | 8 |
| A premature baby may be kept NPO | 89 (52.4) | 81 (47.6) | 4 |
| A premature baby may be given feeding through NGT | 106 (62.4) | 64 (37.6) | 3 |
| Feeding a premature baby through NGT is by pushing milk through a syringe in the tube | 83 (48.8) | 87 (51.2) | 6 |
| Pacifiers may be used for NPO premature baby | 84 (49.4) | 86 (50.6) | 5 |
| Premature babies should receive small and frequent amounts of milk | 120 (70.6) | 50 (29.4) | 1 |
| Burping a premature baby during and after feeding is not essential | 73 (42.9) | 97 (57.1) | 7 |
| Expressed breast milk is best if the baby is unable to breastfeed | 116 (68.2) | 54 (31.8) | 2 |

**Supplementary Table S10.** Phototherapy knowledge items (N=170)

| Item | Correct N (%) | Incorrect N (%) | Rank |
|---|---|---|---|
| The premature baby's temperature should be monitored during phototherapy | 148 (87.1) | 22 (12.9) | 1 |
| The premature baby's position should be changed frequently during phototherapy | 85 (50.0) | 85 (50.0) | 4 |
| The baby's eyes should be covered during phototherapy | 94 (55.3) | 76 (44.7) | 3 |
| The baby's genitalia should be covered during phototherapy | 70 (41.2) | 100 (58.8) | 5 |
| A baby under phototherapy should be kept well hydrated through IV fluid or milk feeding | 118 (69.4) | 52 (30.6) | 2 |

**Supplementary Table S11.** Infection and skin care knowledge items (N=170)

| Item | Correct N (%) | Incorrect N (%) | Rank |
|---|---|---|---|
| The diaper can cover the umbilical cord | 70 (41.2) | 100 (58.8) | 7 |
| The umbilical cord should be kept dry and clean | 160 (94.1) | 10 (5.9) | 3 |
| A soiled diaper should be changed immediately | 164 (96.5) | 6 (3.5) | 1 |
| Hand hygiene should be done before handling a premature baby | 97 (57.1) | 73 (42.9) | 5 |
| The umbilical cord should be covered | 86 (50.6) | 84 (49.4) | 6 |
| Instruments used for the care of the baby could be shared with others | 161 (94.7) | 9 (5.3) | 2 |
| Perineal and umbilical cord areas should be monitored closely for infection | 140 (82.4) | 30 (17.6) | 4 |

**Supplementary Table S12.** Clinical-priority education gaps based on the 10 lowest-scoring knowledge items

| Rank | Domain | Knowledge item | Correct N (%) | Incorrect/gap N (%) | Practical implication |
|---|---|---|---|---|---|
| 1 | Thermoregulation | Premature babies should not be manipulated frequently | 48 (28.2) | 122 (71.8) | Reinforce safe handling and heat-loss prevention |
| 2 | Feeding | Not all premature babies can tolerate breastfeeding | 58 (34.1) | 112 (65.9) | Explain feeding readiness and clinical variation |
| 3 | Phototherapy | The baby's genitalia should be covered during phototherapy | 70 (41.2) | 100 (58.8) | Include phototherapy safety in parent teaching |
| 4 | Infection and skin care | The diaper should not cover the umbilical cord | 70 (41.2) | 100 (58.8) | Reinforce cord care and infection prevention |
| 5 | Feeding | Burping a premature baby during and after feeding is essential | 73 (42.9) | 97 (57.1) | Include practical feeding support before discharge |
| 6 | Feeding | Feeding through NGT should not be done by pushing milk through a syringe | 83 (48.8) | 87 (51.2) | Clarify safe NGT feeding principles |

| | | | | | |
|---|---|---|---|---|---|
| 7 | Feeding | Pacifiers may be used for NPO premature babies | 84 (49.4) | 86 (50.6) | Explain non-nutritive sucking and NPO care |
| 8 | Phototherapy | The baby's position should be changed frequently during phototherapy | 85 (50.0) | 85 (50.0) | Reinforce positioning during phototherapy |
| 9 | Infection and skin care | The umbilical cord should not be covered | 86 (50.6) | 84 (49.4) | Clarify cord exposure and hygiene |
| 10 | Feeding | A premature baby may be kept NPO | 89 (52.4) | 81 (47.6) | Explain why oral feeding may be temporarily withheld |

**Note.** Items are presented from lowest to highest proportion of correct responses. Incorrect responses across these 10 items were counted as clinical-priority education gaps. NGT, nasogastric tube; NPO, nil per os.

**Supplementary Table S13.** Monte Carlo sensitivity checks for selected bivariate associations

| Variable | Pearson $\chi^2$ | Pearson *p*-value | Minimum expected count | Monte Carlo *p*-value | Interpretation |
|---|---|---|---|---|---|
| Previous premature babies | 10.75 | 0.029 | 3.38 | **0.029*** | Significant |
| Number of children, including this baby | 9.25 | 0.055 | 7.96 | 0.054 | Borderline |
| Birth order of index child | 14.68 | 0.023 | 9.16 | **0.022*** | Significant |
| ANC visits group | 10.66 | 0.031 | 11.58 | **0.030*** | Significant |
| Specialist premature-care ANC follow-up | 5.85 | 0.054 | 11.58 | 0.052 | Borderline |
| Prenatal examination by doctor | 5.96 | 0.051 | 15.92 | 0.051 | Borderline |
| Perceived information as adequate | 5.61 | 0.060 | 14.55 | 0.057 | Borderline |
| Mass media during pregnancy | 3.10 | 0.212 | 0.82 | 0.178 | Not significant; sparse |

**Note.** Monte Carlo chi-square sensitivity checks used 200,000 simulated tables. ANC, antenatal care. Borderline refers to *p*-values close to, but not below, 0.05.

**Supplementary Table S14.** Sensitivity binary logistic regression: poor versus good/excellent knowledge

| Predictor | Adjusted OR for poor knowledge | 95% CI | *p*-value |
|---|---|---|---|
| Maternal education level, higher category | 0.88 | 0.56–1.39 | 0.583 |
| Previous premature infants, higher category | 1.45 | 0.77–2.71 | 0.246 |
| Birth order of index child, higher category | 0.90 | 0.65–1.25 | 0.534 |
| Number of ANC visits | 0.98 | 0.87–1.10 | 0.697 |
| No specialist premature-care ANC follow-up, vs yes | 3.07 | 1.11–8.46 | 0.030* |
| No prenatal doctor examination, vs yes | 0.52 | 0.20–1.31 | 0.162 |
| No information after delivery, vs yes | 0.84 | 0.34–2.10 | 0.711 |

**Note.** Outcome was poor knowledge versus good/excellent knowledge. OR >1 indicates higher odds of poor knowledge. ANC, antenatal care; CI, confidence interval; OR, odds ratio.

**Supplementary Table S15.** Pearson correlations between knowledge domains (N=170)

| Domain comparison | Pearson correlation coefficient (r) | *p*-value |
|---|---|---|
| Thermoregulation and feeding | 0.735 | <0.001 |
| Thermoregulation and phototherapy | 0.670 | <0.001 |
| Thermoregulation and infection and skin care | 0.586 | <0.001 |
| Feeding and phototherapy | 0.633 | <0.001 |
| Feeding and infection and skin care | 0.570 | <0.001 |

| Phototherapy and infection and skin care | 0.574 | <0.001 |
|---|---|---|

**Supplementary Table S16.** Additional exploratory and sensitivity analyses

| Predictor | Poor feeding OR (95% CI); *p*-value | Poor phototherapy OR (95% CI); *p*-value | Continuous score beta (95% CI); *p*-value | Clinical-priority gap count IRR (95% CI); *p*-value |
|---|---|---|---|---|
| Maternal education level, higher category | 0.75 (0.48–1.18); 0.212 | 1.05 (0.67–1.63); 0.831 | 0.2 (−5.0 to 5.3); 0.944 | 1.01 (0.92–1.13); 0.780 |
| Previous premature infants, higher category | 1.49 (0.83–2.70); 0.183 | 1.50 (0.82–2.72); 0.186 | **−8.0 (−15.0 to −1.1); 0.024*** | **1.21 (1.06–1.37); 0.004*** |
| Birth order of index child, higher category | 0.91 (0.67–1.25); 0.571 | 1.02 (0.75–1.38); 0.919 | 2.6 (−0.9 to 6.1); 0.142 | 0.96 (0.90–1.03); 0.312 |
| Number of ANC visits | 1.00 (0.90–1.12); 0.982 | 0.97 (0.86–1.08); 0.565 | −0.1 (−1.3 to 1.1); 0.880 | 1.00 (0.97–1.03); 0.998 |
| No specialist premature-care ANC follow-up, vs yes | 1.38 (0.55–3.48); 0.491 | **2.95 (1.14–7.59); 0.025*** | **−11.5 (−21.6 to −1.4); 0.025*** | **1.28 (1.04–1.57); 0.019*** |
| No prenatal doctor examination, vs yes | 0.48 (0.19–1.19); 0.113 | 0.62 (0.25–1.52); 0.300 | 8.7 (−1.4 to 18.8); 0.091 | 0.82 (0.67–1.00); 0.049 |
| No information after delivery, vs yes | 1.21 (0.51–2.91); 0.663 | 0.53 (0.22–1.31); 0.171 | 3.7 (−6.8 to 14.1); 0.492 | 1.00 (0.82–1.22); 0.968 |

**Note.** Poor feeding and poor phototherapy knowledge were defined as domain scores below 50%. The continuous-score model used the overall 30-item knowledge percentage score and standard errors. The clinical-priority gap count was the number of incorrect responses across the 10 lowest-scoring knowledge items and was analysed using Poisson regression with robust standard errors. ANC, antenatal care; CI, confidence interval; IRR, incidence rate ratio; OR, odds ratio.

### Appendix 2. Study questionnaire

#### Interviewer-administered questionnaire (English version)

Please answer the following questions

**Part I: Socio-demographic characteristics**

- Age in years:……..years
- Place of residence: …………………
- Years of education:…………years
- Average family monthly income:……….NIS
- Husband's years of education:……………..years
- Mother's work status: 1- Employed/worker 2- Housewife
- Husband's work status: 1- Employed/worker 2- Not working

**Part II: Obstetric factors**

1. Maternal risk factors

Diabetes mellitus ☐ Pregnancy-induced hypertension ☐ Premature rupture of ☐ membranes

Previous Caesarean section ☐ Twins pregnancy. ☐

☐ Others (specify……………….)

2. Mode of delivery for this baby:

☐ Normal vaginal delivery ☐ Assisted vaginal delivery (vacuum) ☐ Caesarean section

3. No. of previous premature babies:…………………..
4. Gestational age of this baby in weeks:………………..Weeks
5. No. of children, including this baby:……………………..
6. Birth order of this child: 1- First 2- Second 3- Third 4- Fourth or more
7. Sex of this baby: 1- Male 2- Female
8. The birth date of the baby:………………………………

**Part III: Maternal health services**

1- Number of antenatal visits during pregnancy of this baby:……………times

2- Did you receive a special ANC follow-up related to the care of a premature baby? ☐ Yes ☐ No

3- Did you receive a prenatal exam at ANC from a doctor? ☐ Yes ☐ No

4- Did you receive a prenatal exam at ANC from a nurse/midwife? ☐Yes ☐ No

5- Did a healthcare provider tell you how to prepare for preterm delivery?

☐ Yes ☐ No

**Part IV: Source of information**

1- Did you receive information related to the care of a premature baby during pregnancy?

☐ Yes ☐ No

If yes, the source of information was from **(possible to select more than one answer)**:

a. Doctor b. Nurse/midwife c. Mass media d. Relative/friend

2- Did you receive information related to the care of the premature after delivery of this baby?

☐ Yes ☐ No

If yes, the source of information was from **(possible to select more than one answer)**:

a. Doctor b. Nurse/midwife c. Mass media d. Relative/friend

If your answer for the previous 2 questions was no, skip the next question (Q 3)

3- Did you perceive the information you received as adequate? ☐ Yes ☐ No

**Part V: Mother's knowledge regarding care of a premature baby (maximum score 30)**

| A | Thermoregulation | Yes | No |
|---|---|---|---|
| 1. | The premature baby should be placed in a warm environment | / | |
| 2. | The premature baby loses body heat rapidly if uncovered | / | |
| 3. | Covering a premature baby's head helps in keeping the baby's temperature | / | |

| | | | |
|---|---|---|---|
| 4. | Premature babies should be placed near open windows with air drafts | | X |
| 5. | A premature baby could be placed on cold beds or a blanket | | X |
| 6. | Premature babies should be manipulated frequently | | X |
| 7. | The Source of heat is essential to keep a baby's normal temperature | / | |
| 8. | Handling the baby with warmed hands is necessary | / | |
| 9. | Dressing the baby in previously warmed clothes is necessary | / | |
| 10. | The baby should be bathed with warm water with immediate drying and covering after bathing | / | |
| B | Feeding | | |
| 1. | Any premature baby can tolerate breastfeeding | | X |
| 2. | A premature baby may be kept NPO | / | |
| 3. | A premature baby may be given feeding through NGT | / | |
| 4. | Feeding a premature baby through NGT is by pushing milk through a syringe in the tube | | X |
| 5. | Pacifiers may be used for NPO premature baby | / | |
| 6. | Premature babies should receive small and frequent amounts of milk | / | |
| 7. | Burping a premature baby during and after feeding is not essential | | X |
| 8. | Expressed breast milk for baby is best if unable to receive breastfeeding | / | |
| C | Phototherapy | | |
| 1. | The premature baby's temperature should be monitored during phototherapy | / | |
| 2. | The premature baby's position should be changed frequently during phototherapy | / | |
| 3. | The Baby's eyes should be covered during phototherapy | / | |
| 4. | The Baby's genitalia should be covered during phototherapy | / | |
| 5. | Baby under phototherapy should be kept well hydrated through IV fluid or milk feeding | / | |
| D | Infection & Skincare | | |
| 1. | The diaper can cover the umbilical cord | | X |
| 2. | The umbilical cord should be kept dry and clean | / | |
| 3. | A soiled diaper should be changed immediately | / | |
| 4. | Hand hygiene should be done before handling a premature baby | / | |
| 5. | The umbilical cord should be covered | | X |
| 6. | Instruments used for the care of the baby could be shared with others | | X |
| 7. | Perineal and umbilical cord areas should be monitored closely for infection | / | |

**Scoring note.** In Part V, "/" indicates the response scored as correct and "X" indicates the response scored as incorrect; these marks are part of the scoring key and are not participant responses.

#### Interviewer-administered questionnaire (Arabic version)

رقم الاستبانة:.....................

أرجو الإجابة على الأسئلة التالية:

**أولاً: البيانات الديموغرافية والاجتماعية**

| | |
|---|---|
| 1. | **عمرك بالسنوات:**..............................سنة |
| 2. | **مكان السكن (المحافظة):**........................................ |
| 3. | **عدد سنوات تعليمك:**........................................ |
| 4. | **عدد سنوات تعليم الزوج:**........................................ |
| 5. | متوسط الدخل الشهري بالشيقل:....................... |
| 6. | **عمل أم الطفل:** 1- تعمل/موظفة 2- ربة منزل |
| 7. | **عمل والد الطفل:** 1- يعمل/موظف 2- لا يعمل |
| **ثانياً: عوامل متعلقة بالولادة** | |
| 1. | **عوامل خطر عند الأم:** 1- السكر 2. ارتفاع ضغط دم الحمل 3- انفجار مبكر لأغشية الجنين 4- ولادة قيصرية 5- حمل بتوأم 6- أخرى (مع التحديد)......................... |
| 2. | **طريقة ولادة الطفل:** 1- طبيعية مهبلياً 2- مهبلياً باستخدام شفاط 3- قيصرية |
| 3. | عدد الأطفال المولودين خدج سابقاً (غير هذا الطفل): ............ طفل |
| 4. | عمر حمل هذا الطفل بالأسابيع:.....................أسبوع |
| 5. | عدد الأطفال لديك (مع هذا الطفل):................. طفل |
| 6. | **ترتيب هذا الطفل بين أطفالك:** 1- الأول 2- الثاني 3- الثالث 4- الرابع أو أكثر |
| 7. | **جنس الطفل:** 1- ذكر 2- أنثى |
| .8 | تاريخ ميلاد الطفل:............................. |
| **ثالثاً: الخدمات الصحية ذات العلاقة بالأم** | |
| 1. | عدد الزيارات لعيادة الحوامل خلال حملك بهذا الطفل:.................... زيارة |
| 2. | **هل تلقيت متابعة خاصة بعيادة الحوامل متعلقة برعاية طفل خداج:** 1- نعم 2- لا |
| 3. | هل تلقيت فحص من طبيب خلال متابعتك بعيادة الحوامل: 1- نعم 2 – لا |
| 4. | هل تلقيت فحص من ممرضة/قابلة خلال متابعتك بعيادة الحوامل: 1- نعم 2 – لا |
| 5. | هل أخبرك مقدم الخدمة الصحية عن كيفية الإستعداد لولادة طفل مبكراً (خداج): 1- نعم 2 – لا |
| **رابعاً: مصدر المعلومات** | |
| 1. | **هل تلقيت معلومات ذات علاقة برعاية الطفل الخداج خلال فترة الحمل؟** 1- نعم 2 – لا<br>إذا كانت الإجابة بنعم، من كان مصدر تلك المعلومات (يمكنك اختيار أكثر من اجابة حسب ما ينطبق):<br>1- الطبيب 2- الممرضة/القابلة 3- الإعلام 4- أقارب/أصدقاء |

| | | | |
|---|---|---|---|
| 2. | **هل تلقيت معلومات متعلقة برعاية الطفل الخداج بعد ولادة هذا الطفل:**<br>1- نعم 2 – لا<br>إذا كانت الإجابة بنعم، من كان مصدر تلك المعلومات (يمكنك اختيار أكثر من اجابة حسب ما ينطبق):<br>1- الطبيب 2- الممرضة/القابلة 3- الإعلام 4- أقارب/أصدقاء | | |
| 3. | إذا كانت إجابتك للسؤالين السابقين ب (لا) لا تجيبي هذا السؤال:<br>هل تعتبري المعلومات التي تلقيتيها بخصوص رعاية الطفل الخداج كافية؟<br>1- نعم 2 – لا | | |
| **خامساً: معلومات الأم حول العناية بالطفل الخداج** | | | |
| أ | الحفاظ على حرارة الطفل | صحيح | خطأ |
| 1. | الطفل الخداج يجب وضعه في بيئة دافئة | / | |
| 2. | الطفل الخداج يفقد حرارته بسرعة إن لم يغطى | / | |
| 3. | تغطية رأس الطفل الخداج يساعد في الحفاظ على درجة حرارة جسمه | / | |
| 4. | الطفل الخداج يتم وضعه بجانب الشبابيك المفتوحة التي يدخلها تيار هوائي | | X |
| 5. | يمكن وضع الطفل الخداج فوق سرير أو حرام بارد | | X |
| 6. | الأطفال الخدج يتم تحريكهم باستمرار | | X |
| 7. | مصدر للحرارة ضروري بجوار الطفل الخداج للحفاظ على حرارته | / | |
| 8. | تتم ملامسة الطفل الخداج بأيدي دافئة | / | |
| 9. | من الضروري الباس الطفل ملابس تم تدفئتها مسبقاً | / | |
| 10. | تحميم الطفل الخداج يتم بماء دافيء مع سرعة التجفيف وتغطية الطفل بعد الحمام | / | |
| ب | تغذية الطفل | صحيح | خطأ |
| 1. | أي طفل خداج يستطيع ممارسة الرضاعة من ثدي الأم | | X |
| 2. | الطفل الخداج من الممكن ألا يسمح له بأخذ أي شيء عبر الفم | / | |
| 3. | الطفل الخداج من الممكن إعطائه الحليب عبر انبوب يصل للمعدة | / | |
| 4. | تغذية الطفل عبر انبوب المعدة يتم من خلال دفع الحليب بواسطة سرنجة دفعاً في الانبوب | | X |
| 5. | البز الكاذب (اللهاية) يمكن استخدامه مع الطفل الخداج الذي لا يرضع عبر الفم | / | |
| 6. | الطفل الخداج يتم إرضاعه بكميات صغيرة ومتكررة من الحليب | / | |
| 7. | ليس من الضروري تكريع (تشجأة) الطفل الخداج أثناء وبعد الرضاعة | X | |
| 8. | يعتبر حليب الأم المشفوط أفضل للطفل في حال لا يستطيع الرضاعة من ثدي الأم | / | |
| ت | العلاج الضوئي | صحيح | خطأ |
| 1. | يجب مراقبة حرارة الطفل الخداج أثناء وضعه تحت العلاج الضوئي | / | |
| 2. | يجب أن يتم تغيير وضعية الطفل الخداج أثناء وضعه تحت العلاج الضوئي | / | |
| 3. | يجب تغطية عيني الطفل الخداج أثناء وضعه تحت العلاج الضوئي | / | |
| 4. | يجب تغطية الجهاز التناسلي للطفل الخداج أثناء وضعه تحت العلاج الضوئي | / | |

| | | | |
|---|---|---|---|
| 5. | الأطفال تحت العلاج الضوئي يجب الحفاظ على السوائل لديهم من خلال اعطاء الحليب أو السوائل الوريدية | / | |
| ث | العدوى والعناية بالجلد | صحيح | خطأ |
| 1. | الحبل السري يمكن تغطيته بالحفاظة | | X |
| 2. | يجب أن يحافظ على الحبل السري جافا ونظيفاً | / | |
| 3. | الحفاظة المتسخة يجب تغييرها في الحال | / | |
| 4. | غسل الأيدي بالصابون أو معقم ضروري قبل ملامسة الطفل | / | |
| 5. | يجب تغطية الحبل السري | | X |
| 6. | الأدوات المستخدمة للطفل يمكن مشاركتها مع الآخرين | | X |
| 7. | المنطقة التناسلية و الحبل السري يجب مراقبتها باستمرار لعلامات العدوى | / | |
